# Nigral volume loss in prodromal, early, and moderate Parkinson’s disease

**DOI:** 10.1101/2023.08.19.23294281

**Authors:** Jason Langley, Kristy S. Hwang, Daniel E. Huddleston, Xiaoping P. Hu, the Parkinson’s Progression Markers Initiative

## Abstract

The loss of melanized neurons in the substantia nigra pars compacta (SNc) is a hallmark pathology in Parkinson’s disease (PD). Melanized neurons in SNc can be visualized in vivo using magnetization transfer (MT) effects. Nigral volume was extracted in data acquired with a MT-prepared gradient echo sequence in 50 controls, 90 non-manifest carriers (46 LRRK2 and 44 GBA1 nonmanifest carriers), 217 prodromal hyposmic participants, 76 participants with rapid eye movement sleep behavior disorder (RBD), 194 de novo PD patients and 26 moderate PD patients from the Parkinson’s Progressive Markers Initiative. No difference in nigral volume was seen between controls and LRRK2 and GBA1 non-manifest carriers (*F*=0.732; *P*=0.483). A significant main effect in group was observed between controls, prodromal hyposmic participants, RBD participants, and overt PD patients (*F*=9.882; *P*<10^−3^). This study shows that nigral depigmentation can be robustly detected in prodromal and overt PD populations.

## 1. Introduction

Parkinson’s disease (PD) is a heterogeneous neurodegenerative disorder with a variety of motor and non-motor symptoms that can be clinically challenging to diagnose and manage, and there are currently no effective interventions to stop PD neurodegeneration. Empirical evidence suggests that PD-related neurodegeneration starts prior to symptom onset^1-5^ and understanding the magnitude and timing of PD-related neurodegeneration is essential to the development of early-stage diagnostic markers and outcome measures for clinical trials. At risk populations, such as individuals with mutations in the leucine-rich repeat kinase2 (LRRK2) and glucosylceramidase β1 (GBA1) genes^6,7^, patients with rapid eye movement sleep behavior disorder (RBD)^8^ or hyposmia with dopamine transporter deficits^9^, are ideal populations to examine neurodegeneration in the prodromal phase of PD since RBD^10,11^ and hyposmic^12^ patients have a high risk of phenoconverting to PD or other synucleinopathies.

Neuromelanin loss in the substantia nigra pars compacta (SNc) is a hallmark pathology of PD^5,13,14^. The role of SNc in PD pathogenesis has been challenging to study *in vivo* due to a lack of tools to investigate PD-related nigral neurodegeneration in living patients. Incidental magnetization transfer (MT) effects^15^ or explicit MT effects generated by MT preparation pulses^16-18^ can be used to generate neuromelanin-sensitive contrast and delineate melanized structures *in vivo*. Application of MT effects to image depigmentation has revealed PD-related reductions in MT contrast ratios in SNc^15,19-21^, nigral volume^16,22-27^, or the area of SNc in a single slice^28,29^. Nigral regions of interest, derived from images with MT effects, have also been used to examine PD-related microstructural changes^30,31^ or iron deposition^32,33^ in SNc.

In prodromal populations, much of the work using MRI has focused on developing diagnostic markers in populations with RBD. Application of MT effects have found reduced locus coeruleus contrast^34^, SNc area^35^, and SNc volume^36^ in RBD relative to controls. Genetic mutations have not been found to influence cortical thickness or volume of subcortical grey matter structures in LRRK2 and GBA1 non-manifest carriers (NMC)^37^ but changes in nigral iron have been observed in LRRK2 and GBA1 NMC^38,39^. Hyposmic subjects with striatal dopamine transporter (123-I Ioflupane (DaTScan)) deficits have a high risk of phenoconverting^40^ and these results suggest that SNc may also be undergoing neurodegeneration since DaTScan binding ratio is correlated with nigral volume^32^. However, the extent of SNc neuronal loss in hyposmic participants is unknown.

Here, nigral volume is examined in two prodromal populations, one consisting of RBD patients and one consisting of hyposmic participants with dopamine transporter deficits since these populations are highly likely to phenoconvert to PD^40,41^. Nigral volume is examined in a cohort consisting of controls, hyposmic participants with dopamine transporter deficits, RBD, *de novo* PD (early PD, levodopa naïve at study enrollment), and moderate PD from the Parkinson’s Progression Markers Initiative (PPMI). Nigral volumes are also examined in LRRK2 and GBA1 NMC.

## 2. Methods

### 2.1 PPMI Overview

Data used in the preparation of this article were obtained from the Parkinson’s Progression Markers Initiative (PPMI) database. For up-to-date information on the study, visit ppmi-info.org. Full inclusion and exclusion criteria for enrollment in PPMI can be found at www.ppmi-info.org. The PPMI project was approved by the Institutional Review Board or Independent Ethics Committee of all participating sites in Europe, including National and Kapodistrian University of Athens (Greece), Hospital Clinic de Barcelona and Hospital Universitario Donostia (Spain), Innsbruck University (Austria), University of Marburg (Germany), Imperial College London (United Kingdom), University of Salerno (Italy), Pitié-Salpêtrière Hospital (France), and in the United States of America, including Emory University, Johns Hopkins University, University of Alabama at Birmingham, PD and Movement Disorders Center of Boca Raton, Boston University, Northwestern University, University of Cincinnati, Cleveland Clinic Foundation, Baylor College of Medicine, Institute for Neurodegenerative Disorders, Columbia University Medical Center, Beth Israel Medical Center, University of Pennsylvania, Oregon Health and Science University, University of Rochester, University of California at San Diego, and University of California, San Francisco. Each participant provided informed, written consent, prior to enrolling in PPMI.

Each participant enrolled in PPMI underwent cognitive testing, a physical exam, and genetic testing for pathogenic variants of the LRRK2 (G2019S) or GBA1 (N409S, R535H, L29Afs*18, L483P) genes. The Movement Disorders Society Unified Parkinson’s Disease Rating Scale (MDS UPDRS)^42^, performed by a movement disorders neurologist certified in the use of this scale, was used to measure motor and nonmotor parkinsonian symptoms at each visit. In addition, each participant underwent the PPMI cognitive battery including the Montreal Cognitive Assessment (MoCA) at each visit^43^. Olfactory function was assessed by the University of Pennsylvania Smell Identification Test (UPSIT) at baseline.

### 2.2 Participants

Criteria for inclusion of subjects from the PPMI database used in this analysis were as follows: 1) participants must be scanned with a MT prepared GRE sequence on a Siemens scanner. A total of 653 participants (50 controls, 46 LRRK2 NMC, 44 GBA1 NMC, 76 RBD, 217 prodromal hyposmic participants, 194 *de novo* PD patients, and 26 moderate PD patients (PD patients with LRRK2 or GBA1 mutations at the 48-month time point) met these criteria. NMC LRRK2 and GBA1 mutations were confirmed to have pathogenic variants of LRRK2 (G2019s) or GBA1 (N409S, R535H, L29Afs*18, L483P) genes. These participants were included in the analysis if they had MDS-UPDRS-III scores ≤ 5 at the 48-month time point and were not diagnosed with PD. Non-manifest LRRK2 and GBA1 participants were taken at the 48-month time point since that was the first time point containing MT-prepared GRE images. All hyposmic participants used in the analysis had hyposmia based on the UPSIT, dopamine transporter deficits, and were not diagnosed with PD. The moderate PD patients are not the same individuals from the *de novo* PD group. Imaging data were downloaded between July 2022 and December 2023.

### 2.3 MRI Acquisition

MRI data used in this analysis were acquired on Siemens MRI scanners. NM-MRI data were acquired using a 2D MT-prepared GRE sequence^17,18^: mean/min/max echo time (TE)=4.12 ms / 2.88 ms / 5 ms, mean/min/max repetition time (TR) = 478 ms / 450 ms / 691 ms, slice thickness 2 mm, in plane resolution 0.5×0.5 mm^2^, mean/min/max flip angle (FA) = 39.7°/22°/40°, mean/min/max bandwidth = 464 Hz/pixel / 122 Hz/pixel / 507 Hz/pixel, 16 contiguous slices, and magnetization transfer preparation pulse (300°, 1.2 kHz off resonance, 10 ms duration), 5 or 10 measurements. A T_1_ magnetization-prepared rapid gradient echo (MP-RAGE) sequence was acquired with the following parameters: TE/TR= 2.62 ms/2300 ms, inversion time = 900 ms, FA=9°, voxel size = 1.0 × 1.0 × 1.0 mm^3^ and used to derive a transform between Montreal Neurological Institute (MNI) common space and native T_1_-weighted images.

### 2.4 Image Processing

MRI data was processed using the FMRIB Software Library (FSL). A transformation was derived between each individual’s T_1_-weighted image and 2 mm Montreal Neurological Institute (MNI) T_1_-space using FMRIB’s Linear Image Registration Tool (FLIRT) and FMRIB’s Nonlinear Image Registration Tool (FNIRT) in the FSL software package using the following steps^44,45^. The T_1_-weighted image was brain extracted using the brain extraction tool (BET). Next, an affine transform was used to align the brain extracted T_1_-weighted images with the MNI brain extracted image. Finally, a nonlinear transformation was used to generate a transformation from individual T_1_-weighted images to T_1_-weighted MNI T_1_-space.

For each participant, individual MT-prepared GRE measurements were corrected for motion by registering all the measurements to the first measurement using a rigid-body transform in FLIRT, denoised,^46^ and then averaged. Finally, a transform was derived between each individual’s T_1_-weighted image and the averaged MT-prepared GRE image with a boundary-based registration cost function. This transform was then inverted. This procedure is illustrated in **Figure 1**.

**Figure 1.**
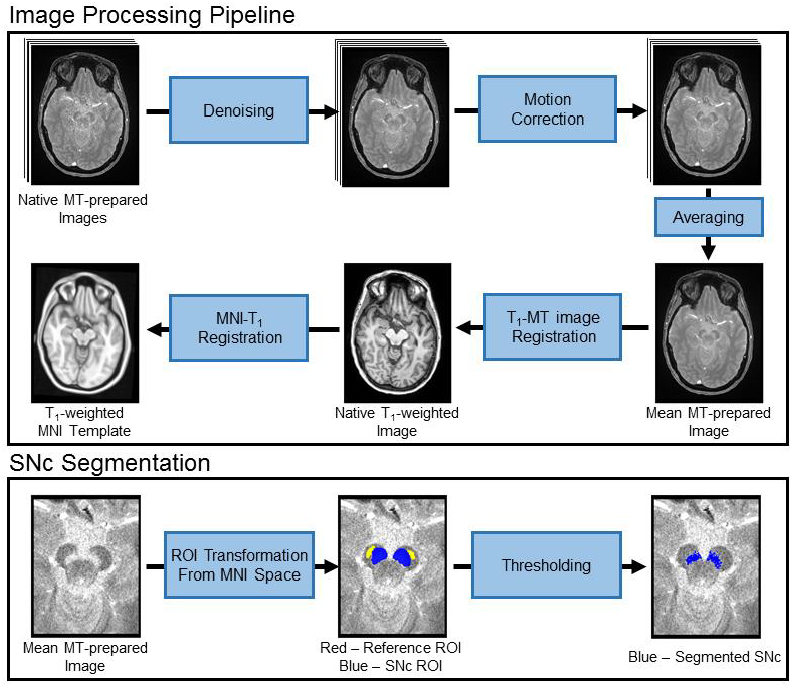
Schematics illustrating the processing steps for the MT-prepared GRE data and SNc segmentation procedure.

SNc volume was segmented in native space using an automated thresholding method. To ensure consistent placement of reference regions of interest (ROIs), a reference ROI in the cerebral peduncle was created using the MNI template and, for each subject, the cerebral peduncle ROI was transformed to individual MT-prepared GRE images using the MNI-T_1_ and T_1_-GRE transforms described in previous paragraphs. The transform was done in a single step to reduce interpolation. The use of standard space ROIs ensured that the reference ROI was placed in similar locations for each subject. The mean (denoted μ_ref_), and standard deviation (σ_ref_) of the signal intensities were measured in the reference ROI.

Next, a standard space SNc atlas was used to localize regions surrounding SNc for thresholding.^47^ This atlas was thresholded at a level of 5%, dilated, and transformed from standard space to individual MT-prepared gradient echo images. The ROIs for thresholding were dilated to ensure that the entire SNc was included for thresholding. Voxels in the resulting ROIs with intensity >μ_ref_+2.8σ_ref_ were considered to be part of SNc. This procedure is illustrated in **Figure 1**.

### 2.5 Statistical Analysis

All statistical analyses were performed using IBM SPSS Statistics software version 28 (IBM Corporation, Somers, NY, USA) and results are reported as mean ± standard deviation. A *P* value of 0.05 was considered significant for all statistical tests performed in this work. Normality of SNc volume was assessed using the Shapiro-Wilk test for each group and all data was found to be normal.

For demographic data, analysis of variance (ANOVA) was used to assess differences in age, years of education, MDS UPDRS-III OFF score, and MoCA of the prodromal PD, overt PD (*de novo*, moderate), and control groups. Chi square was used to examine differences in sex between groups.

The effect of genetic mutations (GBA1, LRRK2) in NMC and controls on SNc volume was assessed with an analysis of covariance (ANCOVA) analysis controlling for age, total brain volume, imaging site, and sex.

The effect of group (control, RBD, hyposmia, *de novo* PD, moderate PD) was tested with an ANCOVA for SNc volume controlling for sex, age, total brain volume, and imaging site. For all ANCOVAs, if the interaction was significant, post hoc comparisons between each pair of groups were performed using respective two-tailed t-tests.

The effect of nigral volume on clinical measures (MDS UPDRS-III OFF score, disease duration) was assessed by correlating nigral volume with clinical measures in the combined PD group (*de novo*+moderate). Correlations between clinical measures and nigral volume were performed using Pearson correlations in PD groups, controlling for age and total brain volume.

## 3. Results

### 3.1 Sample Demographics

A total of 140 participants were used in the analysis of nigral volume in non-manifest LRRK2 (46 participants), non-manifest GBA1 (44 participants), and control (50 participants) groups. No differences in age (*F*=1.505; *P*=0.226), MoCA (*F*=0.805; *P*=0.449), MDS UPDRS-III score (*F*=1.138; *P*=0.324) or education (*F=*2.270; *P*=0.107) were observed between LRRK2 NMCs, GBA1 NMCs, and the control group without genetic mutations. A significant difference in sex was found between the LRRK2 NMC and the control group (χ^2^=4.112; *P=*0.043) with the control group having more males than the LRRK2 NMC group. Demographic information for the non-manifest LRRK2 and GBA1 analysis is summarized in **Table 1**.

**Table 1.** Demographic information for the analysis examining effect of genetic mutation on nigral volume in non-manifest participants. Data is presented as mean ± standard deviation unless noted otherwise. ANOVAs were used for group comparisons of age, education, UPDRS-III, and MoCA from which *P* values are shown. CO – control; MDS UPDRS-III – Movement Disorders Society Unified Parkinson’s Disease Rating Scale Part III; MoCA - Montreal Cognitive Assessment.

|  | CO<br>(N=50) | Non-manifest Carriers |  | P |
| --- | --- | --- | --- | --- |
|  |  | LRRK2<br>(N=46) | GBA1<br>(N=44) |  |
| Sex [M/F] | [31/19] | [19/27] | [19/25] | **>0.043** |
| Age [years] | 62.4 ± 11.9 | 65.3 ± 6.8 | 64.8 ± 6.0 | 0.226 |
| Education [years] | 17.2 ± 3.5 | 17.5 ± 2.3 | 18.4 ± 2.5 | 0.107 |
| MoCA | 27.7 ± 2.0 | 28.0 ± 1.6 | 27.5 ± 2.1 | 0.449 |
| MDS UPDRS-III | 1.7 ± 2.1 | 2.0 ± 2.5 | 2.5 ± 2.4 | 0.324 |

A total of 563 participants were used in the analysis of nigral volume in control (50 participants), hyposmia (217 participants), RBD (76 participants), *de novo* PD (194 participants), and moderate PD (26 participants) groups. A significant difference in sex was observed between the hyposmic group and the control (χ^2^=10.158; *P*=0.001), RBD (χ^2^=32.052; *P*<10^−3^), and *de novo* PD (χ^2^=26.770; *P*<10^−3^) groups as well as between the RBD group and the moderate PD group (χ^2^=4.985; *P*=0.026). Comparisons of sex were not significant between other groups (*Ps*>0.058). A significant difference in age (*F*=13.065; *P*<10^−3^) was seen between the groups with the control group being younger, on average, as compared to the RBD (*P*<10^−3^), hyposmia (*P*<10^−3^), and moderate PD (*P*=0.001) groups. The RBD (*P*<10^−3^), hyposmia (*P*<10^−3^), and moderate PD (*P*=0.002) groups were older, on average as compared to the *de novo* PD group. No difference in age was seen between the RBD, hyposmia, or moderate PD groups (*P*s>0.332). A significant difference was found in MoCA (*F*=2.006; *P*=0.114) with higher MoCA scores seen in the control group relative to the hyposmia (*P*=0.011), RBD (*P*=0.005), and the moderate PD (*P*=0.009) groups. A significant difference was observed in MDS UPDRS-III OFF score (*F*=230.820; *P*<10^−3^) with higher MDS UPDRS-III scores seen in the moderate PD group relative to the hyposmia, RBD, *de novo* PD, and control groups (*P*<0.008). Higher MDS UPDRS-III scores were seen in the *de novo* PD group as compared to the hyposmia, RBD, and control groups (*Ps<*10^−3^). The hyposmia and RBD group exhibited higher MDS UPRDS-III scores relative to the control group (*Ps<*0.003). A significant difference was seen in UPSIT score between the groups (*F=*18.736; *P<*10^−3^) with the hyposmic (*P<*10^−3^), RBD (*P<*10^−3^), and *de novo* PD (*P<*10^−3^) groups showing reduced olfactory function relative to controls. No difference in education (*F*=0.077; *P*=0.989) was seen between the groups. Demographic information for the control, hyposmia, RBD, *de novo* PD, and moderate PD groups is summarized in **Table 2**.

**Table 2.** Demographic information for the groups used in the Control-PD pathology analysis. Data is presented as mean ± standard deviation unless noted otherwise. ANOVAs were used for group comparisons of age, education, UPDRS-III, and MoCA from which *P* values are shown. MDS UPDRS-III was measured in the OFF state. UPSIT scores were not released for the moderate PD participants. CO – control; MoCA - Montreal Cognitive Assessment; MDS UPDRS-III – Movement Disorders Society Unified Parkinson’s Disease Rating Scale Part III; RBD – rapid eye movement sleep behavior disorder; UPSIT – University of Pennsylvania Smell Identification Test.

|  | CO<br>(N=50) | RBD<br>(N=76) | Hyposmia<br>(N=217) | *de novo* PD<br>(N=194) | Moderate PD<br>(N=26) | P |
| --- | --- | --- | --- | --- | --- | --- |
| Sex [M/F] | [31/19] | [57/19] | [81/136] | [122/72] | [14/12] | *P*<sub>s</sub>**>10<sup>-3</sup>** |
| Age [years] | 62.4 ± 11.9 | 67.5 ± 5.4 | 68.5 ± 5.7 | 63.0 ± 10.0 | 68.0 ± 7.6 | **<10<sup>-3</sup>** |
| LRRK2/GBA1/None | 0/0/33 | 0/0/76 | 0/0/217 | 5/1/188 | 9/17/0 | – |
| Disease Duration [years] | – | – | – | 1.0 ± 0.9 | 6.7 ± 2.1 | **<10<sup>-3</sup>** |
| Levodopa Equivalents | – | – | – | 12.4 ± 67.9 | 698.3 ± 454.1 | **<10<sup>-3</sup>** |
| MDS UPDRS-III OFF score | 1.7 ± 2.1 | 4.1 ± 4.0 | 5.1 ± 5.4 | 23.2 ± 9.7 | 28.9 ± 11.9 | **<10<sup>-3</sup>** |
| Hoehn & Yahr | 0.0 ± 0.0 | 0.0 ± 0.0 | 0.0 ± 0.0 | 1.7 ± 0.5 | 2.1 ± 0.4 | **<10<sup>-3</sup>** |
| UPSIT | 34.6 ± 4.1 | 23.6 ± 7.8 | 23.2 ± 7.4 | 23.7 ± 7.7 | – | **<10<sup>-3</sup>** |
| Education [years] | 16.9 ± 3.4 | 16.8 ± 2.8 | 16.8 ± 2.8 | 16.8 ± 3.2 | 16.3 ± 3.6 | 0.989 |
| MoCA | 27.7 ± 2.0 | 26.5 ± 2.7 | 26.8 ± 2.1 | 27.2 ± 2.3 | 26.1 ± 2.9 | **0.009** |

Twenty-six moderate PD participants (17 LRRK2 participants; 9 GBA1 participants) were used to assess the impact of genetic mutation on nigral volume in overt PD. No group differences in age (*F*=2.547; *P*=0.124), sex (χ^2^=0.490; *P*=0.484), disease duration (*F*=1.199; *P*=0.284), levodopa equivalents (*F*=0.034; *P*=0.856), MDS UPDRS-III score (*F*=0.732; *P*=0.403), Hoehn and Yahr score (*F*=0.426; *P*=0.521) or MoCA (*F*=0.173; *P*=0.681) were observed between moderate PD patients with LRRK2 and GBA1 mutations. Demographic information for the manifest LRRK2 and manifest GBA1 moderate PD groups is summarized in **Table 3**.

**Table 3.** Demographic information for the analysis examining effect of genetic mutation on nigral volume in manifest PD participants at the 48-month time point. Data is presented as mean ± standard deviation unless noted otherwise. ANOVAs were used for group comparisons of age, education, UPDRS-III, and MoCA from which *P* values are shown. CO – control; MDS UPDRS-III – Movement Disorders Society Unified Parkinson’s Disease Rating Scale Part III; MoCA - Montreal Cognitive Assessment.

|  | Moderate PD |  |  |
| --- | --- | --- | --- |
|  | LRRK2 Carrier | GBA1 Carrier |  |
| Sex [M/F] | [10/7] | [4/5] | 0.484 |
| Age [years] | 66.1 ± 7.3 | 71.0 ± 7.6 | 0.124 |
| Disease Duration [years] | 6.4 ± 1.8 | 7.2 ± 2.2 | 0.284 |
| Levodopa Equivalents | 710.4 ± 366.6 | 675.4 ± 612.0 | 0.856 |
| MDS UPDRS-III | 30.3 ± 9.7 | 25.3 ± 16.6 | 0.403 |
| Hoehn & Yahr | 2.1 ± 0.5 | 2.0 ± 0.0 | 0.521 |
| MoCA | 25.9 ± 3.0 | 26.4 ± 2.8 | 0.449 |
| Education [years] | 16.4 ± 4.0 | 16.2 ± 2.8 | 0.931 |

### 3.2 Non-manifest Comparisons

The effect of LRRK2, GBA1, and no genetic mutation on nigral volume in nonmanifest carriers and controls was tested with an ANCOVA analysis with the number of measurements in the acquisition protocol, total brain volume, sex, and age as covariates. ANCOVA analysis revealed no difference in nigral volume between NMCs of LRRK2 (381 mm^3^ ± 102 mm^3^) and GBA1 (379 mm^3^ ± 83 mm^3^) mutations and control participants (*F*=0.732; *P*=0.483). Total brain volume (*F*=1.454; *P*=0.230) and site (*F*=0.263; *P*=0.609) were not significant covariates in the model. Age (*F*=6.208; *P*=0.014) and sex (*F*=6.485; *P*=0.012) were significant covariates in the model (*F*=6.485; *P*=0.012) with reduced nigral volume associated with older age and male participants. These comparisons are shown in **Figure 2** and a spatial comparison of mean population SNc volume in the LRRK2 NMCs, GBA1 NMCs, and controls is shown in **Figure 3**.

**Figure 2.**
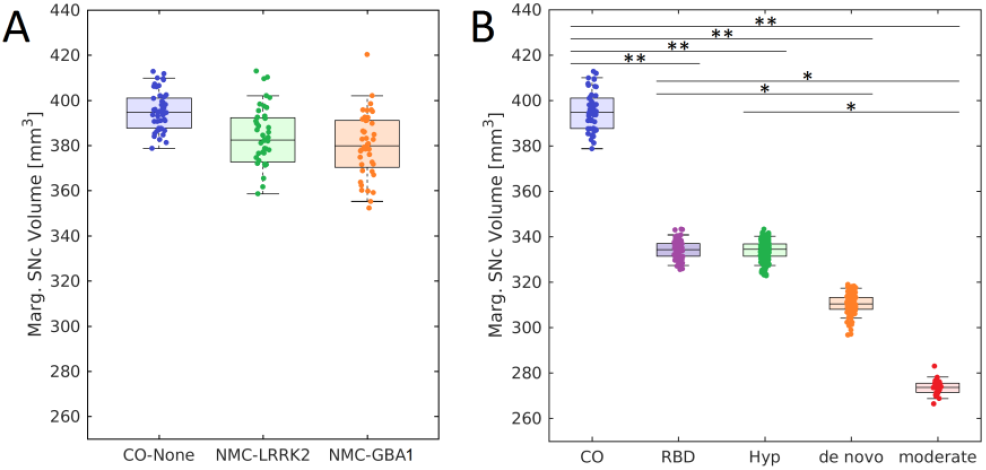
Comparisons of nigral volume marginal means in LRRK2 NMC, GBA1 NMC, and non-carrier controls are shown in A. Comparisons of nigral volume marginal means in the control cohort, RBD, hyposmia, *de novo* PD cohort, and moderate PD cohort is shown in B. In both box plots, the top and bottom of the box denote the 25^th^ and 75^th^ percentiles, respectively, with the line denoting the median value. * and ** denote significant levels of *P*<0.05, *P*<0.01, respectively.

**Figure 3.**
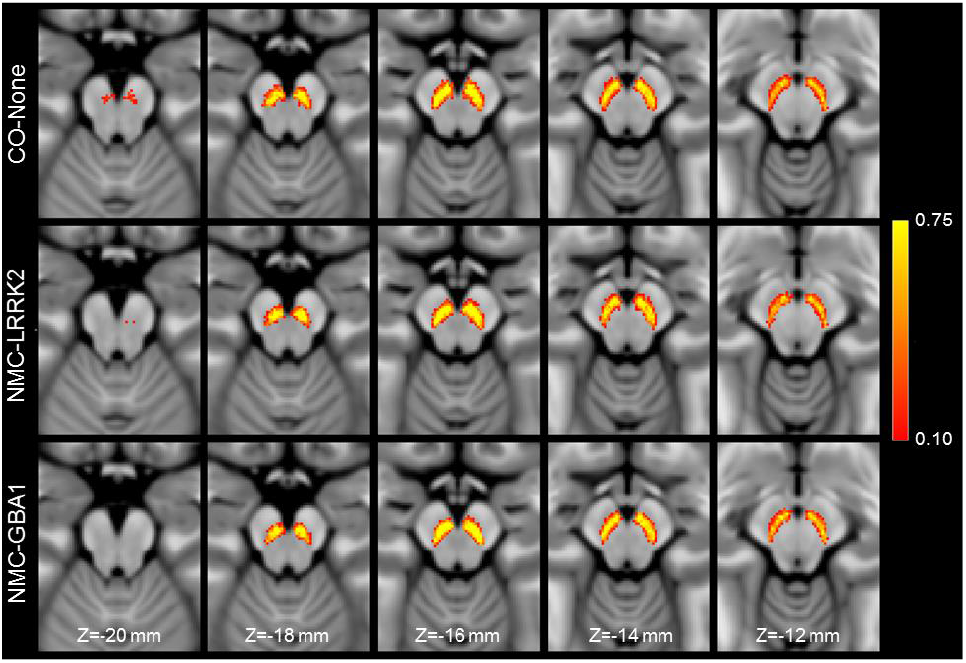
A comparison of SNc population mean volume in the control group (top row), non-manifest LRRK2 carriers (middle row), and non-manifest GBA1 carriers (bottom row). For each group, the SNc population mean volume was created by transforming SNc masks from individual participants to MNI space and then averaging.

### 3.2 Nigral Volume Group Comparisons

**Figure 4** shows a comparison of mean MTC images in the control group (noncarriers), hyposmia group, RBD group, *de novo* PD group, and moderate PD group. The effect of group (control, hyposmia, RBD, *de novo* PD, moderate PD) on SNc volume was assessed using an ANCOVA with number of measurements in the site, age, total brain volume, and sex as covariates. A significant main effect of group (*F*=9.882; *P*<10^−3^) revealed reduced SNc volume in the hyposmia (*P*<10^−3^), RBD (*P*=0.002), *de novo* PD (*P*<10^−3^), and moderate PD (*P*<10^−3^) groups relative to the control group. The moderate PD group showed reduced nigral volume as compared to the hyposmia (*P*=0.012) and RBD (*P*=0.004) groups. A significant difference was seen between the RBD group and the *de novo* PD group (*P*=0.025). No other comparisons were significant (*Ps*>0.065). Sex was a significant covariate in the model (*F*=17.170; *P<*10^−3^) with reduced nigral volume associated with male participants. Site (*F*=3.385; *P=*0.066), age (*F*=0.005; *P=*0.944), and total brain volume (*F*=3.233; *P=*0.073) were not significant covariates. A comparison of SNc population mean volumes is shown in **Figure 5** and marginal means for each group are summarized in **Table 4**.

The effect of genetic mutation (GBA1, LRRK2) on SNc volume in the moderate PD group was tested using an ANCOVA with the number of measures in the NM protocol and sex as covariates. No main effect of group (*F*=0.822; *P*=0.496) was seen in SNc volume (GBA1: 259 mm^3^ ± 113 mm^3^; LRRK2: 262 mm^3^ ± 106 mm^3^). Sex (*F*=0.910; *P*=0.351) and site (*F*=1.624; *P*=0.641) were not significant covariates in the model. Reduced SNc volume was seen in moderate PD patients with the LRRK2 mutation as compared to nonmanifest LRRK2 carriers (NMC: 381 mm^3^ ± 102 mm^3^; *F*=5.997; *P*=0.017). Similarly, reduced SNc volume was observed in moderate PD patients with GBA1 mutation as compared to nonmanifest GBA1 carriers (NMC: 379 mm^3^ ± 83 mm^3^; *F*=15.825; *P*<10^−3^).

**Table 4.** Structure volumes from the marginal means in the control-pathology analysis. Data is presented as mean ± standard deviation. ANCOVAs were used for group comparisons of SNc volume from which the *P*-values and *F*-values are shown.

|  | Control | Hyposmia | RBD | *de novo* PD | Moderate PD | *F* | *P* |
| --- | --- | --- | --- | --- | --- | --- | --- |
| SNC Volume | 395.2 ± 117.0 | 332.1 ± 100.7 | 336.0 ± 96.6 | 309.4 ± 100.0 | 291.2 ± 118.3 | 9.882 | <10<sup>-3</sup> |

**Figure 4.**
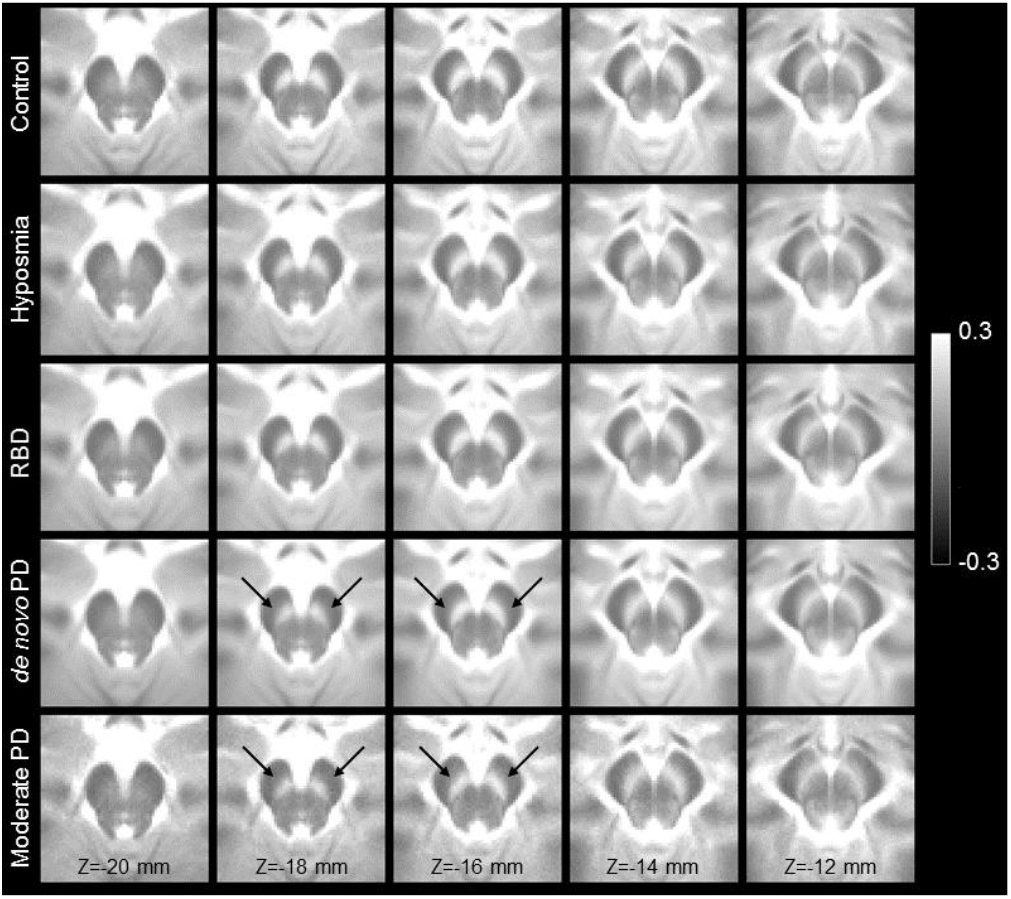
A comparison of mean SNc contrast in control (top row), hyposmia (second row), RBD (third row), *de novo* PD (fourth row), and moderate PD (bottom row) groups. Reduced contrast can be seen in the prodromal and PD groups as compared to the controls in slices Z=−18 mm and Z=−16 mm. For each group, the mean magnetization transfer contrast (MTC) image was created by transforming MTC images from individual participants to MNI space and then averaging. Arrows point to the regions exhibiting a loss of contrast in the de novo PD and moderate PD groups as compared to controls.

**Figure 5.**
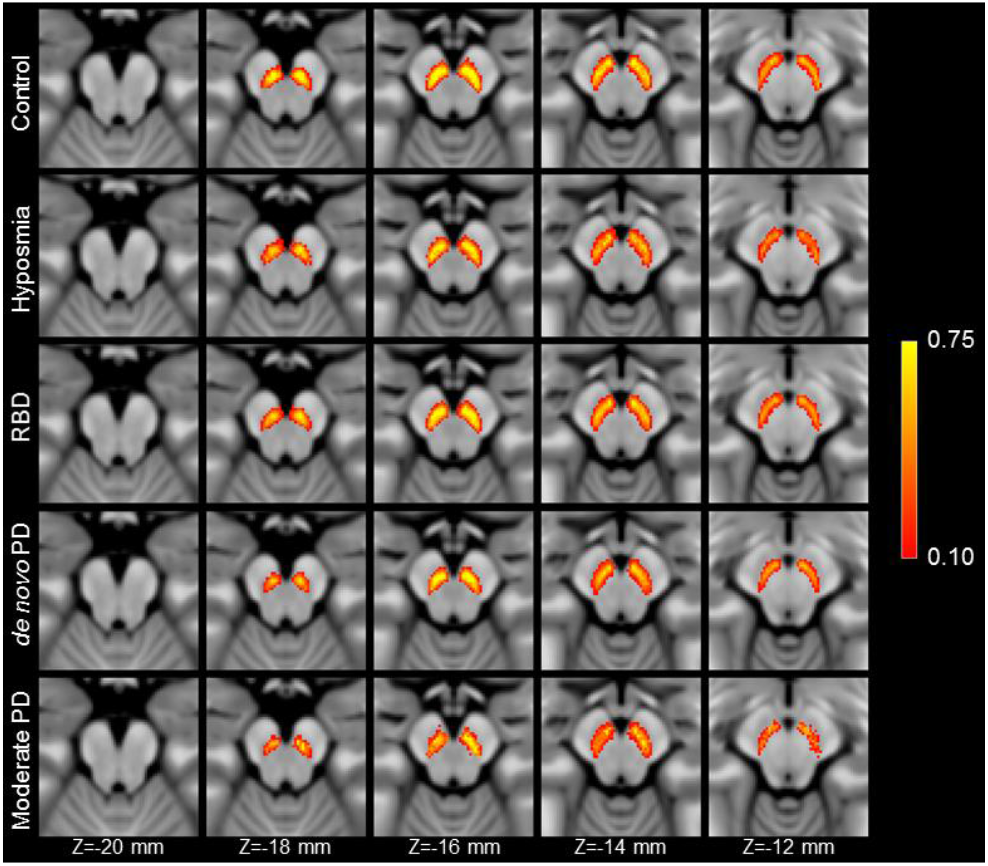
A comparison of SNc population means in control (top row), hyposmia (second row), RBD (third row), *de novo* PD (fourth row), and moderate PD (bottom row) groups. Reduced volume can be seen in all pathologic groups as compared to the controls in slices Z=−18 mm and Z=−16 mm. For each group, the SNc population mean was created by transforming SNc masks from individual participants to MNI space and then averaging.

### 3.2 Clinical Correlations

The effect of disease severity (MDS UPDRS-III OFF score) and disease duration on nigral volume was first assessed, separately, with Pearson correlations controlling for age and sex, in the *de novo* and moderate PD groups. A significant negative correlation was seen between nigral volume and disease duration (*r*=−0.176; *P*=0.019) but no association was seen between MDS UPDRS-III OFF score and nigral volume (*r*=0.075, *P*=0.322) in the *de novo* PD group. No associations between nigral volume and MDS UPDRS-III OFF score (*r*=−0.276; *P*=0.253) or nigral volume and disease duration (*r*=−0.280; *P*=0.246) were observed in the moderate PD group. Pearson correlations, controlling for age and sex, in the combined (*de novo*+48 month) PD group yielded a significant correlation between nigral volume and disease duration (*r*=−0.222, *P*=0.001) with longer disease duration correlated with lower nigral volume. No association was observed between MDS UPDRS-III OFF score and nigral volume (*r*=−0.008; *P*=0.914).

The relationship between UPSIT score and nigral volume was assessed with Pearson correlations controlling for age and sex in the hyposmia prodromal group, RBD group, and *de novo* PD group separately. No associations were observed between nigral volume and UPSIT in the *de novo* PD group (*r*=−0.080; *P*=0.284), RBD group (*r*=−0.039; *P*=0.745), or hyposmia group (*r*=−0.071; *P*=0.302).

## 4. Discussion

This study examined PD-related SNc degeneration in prodromal participants (hyposmic and RBD), *de novo* PD participants, and moderate PD participants. Standard space ROIs were used to define SNc regions used in the thresholding-based segmentation procedure and this method has been shown to exhibit high scan-rescan reproducibility^48-50^. Application of the method found no difference in nigral volume between NMC of LRRK2 and GBA1 mutations and controls. Significant volume loss was seen in SNc of the prodromal PD groups (hyposmia, RBD) as well as in both PD groups (*de novo*, moderate) as compared to controls. In addition, nigral volume in the moderate PD group was reduced as compared to the prodromal groups. Finally, SNc volume was similar for PD patients with LRRK2 and GBA1 genetic mutations in the moderate PD group.

A prior study examining striatal binding ratio from dopamine transporter imaging (123-I Ioflupane DaTScan) in LRRK2 and GBA1 NMCs in the PPMI dataset did not observe DaTScan binding deficits in GBA1 and LRRK2 NMCs at baseline^51^. In addition, no progression was seen in striatal binding ratio in LRRK2 NMCs over two years^52^. As DaTScan striatal binding ratio is correlated with nigral volume^32^, these results suggest that GBA1 and LRRK2 NMCs will have similar nigral volume as controls. Our analysis revealed no difference in nigral between controls and GBA1 and LRRK2 NMCs at the 48-month time point. At the 24-month time point, only 5 of 175 of the LRRK2 NMC participants in PPMI phenoconverted to PD^52^. Similarly, only 3 GBA1 NMC participants with MT-prepared GRE images phenoconverted to PD to date. Thus, it is possible that the lack of a significance difference in nigral volume between NMC and controls may be due to incomplete penetrance of the LRRK2 and GBA1 mutations. However, we do not yet know the lifetime likelihood of phenoconversion to PD in the NMCs in this study, and the sample may also contain many NMCs who are a decade or more away from phenoconversion to PD. Once phenoconversion has occurred in a larger number of NMCs it will be important to study SNc volume as a predictor of phenoconversion.

Olfactory dysfunction is a common symptom of PD^53^ and may precede clinical diagnosis by at least 4 years^54-56^. The prodromal hyposmic participants used here have dopamine transporter deficits and these participants are highly likely to phenoconvert to PD^40,41^. Dopamine transporter deficits suggest this population is experiencing nigral volume loss^32^. Consistent with this posit, reduced nigral volume was observed in the prodromal hyposmic group as compared to controls. Taken together, these results suggest the nigrostriatal system is undergoing neurodegeneration in the prodromal hyposmic group.

Patients who have RBD and no other neurological condition are highly likely to develop an overt synucleinopathy (PD, dementia with Lewy bodies, multiple system atrophy) later in life^57-59^, and RBD is considered to be a prodromal stage of alpha synucleinopathies. Imaging studies have reported reductions in nigral volume or contrast ^35,36,60^ in RBD patients relative to controls. In agreement with these studies, we observed a reduction in nigral volume of the RBD group relative to the control group and a further reduction in nigral volume was observed in the PD group relative to the RBD group.

Using a lateral-ventral SNc ROI^21^, reductions in the proportion of individuals sharing a voxel in the group SNc atlases were found to be reduced in both PD groups relative to controls (controls = 0.38; *de novo*= 0.27; moderate PD = 0.25). The reduction in the proportion of individuals sharing a voxel in these areas agrees with prior studies which found reductions in nigral width^29,61^, loss of contrast in the posterior portion of SNc^27^, or a loss of contrast in the lateral-ventral portions of SNc^21^. Further, these regions have been shown to overlap with nigrosome-1, the subregion of SNc with the greatest loss of melanized neurons^62,63^, and loss of contrast in these regions may be due to depletion of melanized neurons in nigrosome-1.

In contrast to an earlier study^27^, no correlation was seen between nigral volume and MDS-UPDRS-III score in the *de novo* population or in the combined (*de novo* + moderate) PD population. This lack of correlation may be due to an absence of PD patients with more severe motor symptoms in our PD populations. PD populations studied in prior work included PD patients with a wide range of disease severity and disease duration^27^.

Nigral volume was negatively associated with disease duration in the *de novo* PD group alone as well as in the combined PD group (*de novo* + moderate). These results suggests that loss of nigral neurons continues as PD progresses from the *de novo* stage into moderate stage of PD and agree with earlier studies that found nigral volume is negatively associated with disease duration^25,26^. However, results examining nigral volume in the moderate PD group should be interpreted with caution since these participants are LRRK2 and GBA1 carriers and the etiology of PD in these populations may be different than that of idiopathic PD. Larger multi-contrast longitudinal imaging studies examining the effect of genotype on nigral characteristics in PD are needed to elucidate changes in SNc in genetic and idiopathic PD populations.

The current findings provide additional evidence that MT-effects robustly detect nigral depigmentation in prodromal populations PD (hyposmic and RBD), *de novo* PD, and moderate PD groups. The moderate (48-month time point) PD group experienced greater PD-related nigral volume loss as compared to the prodromal and *de novo* PD groups. Genetic mutation was not found to influence nigral volume in NMCs.

## Acknowledgements

PPMI – a public-private partnership – is funded by the Michael J. Fox Foundation for Parkinson’s Research and funding partners, including [list the full names of all of the PPMI funding partners found at www.ppmi-info.org/about-ppmi/who-we-are/study-sponsors].

This work is supported by the NIH-NINDS 1K23NS105944-01A1 (Huddleston), NIH-NIA 1U19AG071754-01 (Huddleston, Hu, Langley), the Department of Veteran Affairs 1I01RX002967-01A2 (Huddleston), the Emory American Parkinson’s Disease Association Center for Advanced Research (Huddleston), the Emory Lewy Body Dementia Association Research Center of Excellence (Huddleston), and the Michael J Fox Foundation (MJF-10854, MJFF-010556; Huddleston, Hu, Langley).

## Data Availability Statement

The data that support the findings of this study are available from the PPMI database (https://www.ppmi-info.org/access-data-specimens/data).

